# Effect of ethanol cleaning on the permeability of FFP2 masks

**DOI:** 10.1101/2020.04.28.20083840

**Authors:** Roland Lenormand, Guillaume Lenormand

## Abstract

In this study we assessed the effect of ethanol on the filtering properties of FFP2 masks. The permeability of parts of a FFP2 mask was measured before and after six cleanings with ethanol. As for any porous medium, the filtering properties of a mask are related to the size and tortuosity of the pores of the filter, and are quantified by its permeability. Any damage to the filter will change its permeability. We show here that after six cleaning cycles, the permeability remains very close to the permeability before cleaning. Amid the COVID-19 pandemic and the shortage of protective masks, this study suggests that ethanol could be used to sanitize a FFP2 mask without significantly altering its filtering properties. Additional measurements on FFP2 and N95 masks from different manufacturers need to be performed to validate this study.

## Introduction

Amid the COVID-19 pandemic and the shortage of protective masks, many essential workers have had to rely on a limited number of protective masks. As the pandemic progresses, portions of the population have been asked to wear masks while in public spaces (1), thereby increasing the demand for protective masks. These masks are made of polyester and other synthetic fibers, acting as a filter to capture droplets while inhaling and exhaling. Importantly they are designed for single use.

In this study, we asked the following question: does the sanitization of a FFP2 mask with 99% ethyl alcohol alter its filtering properties? We hypothesize that the sanitization of a mask with ethyl alcohol between uses would reduce transmission of pathogens such as the SARS-CoV-2 virus.

Like any porous media, the filtering property of a mask is related to the size and tortuosity of the pores of the filter, a property that can be quantified by a permeability measurement. The permeability of a porous media is a measure of the ability of a fluid to pass through it (2). A highly permeable material provides low resistance to the fluid passing through it, whereas a material with low permeability provides more resistance to flow. If the porous media, such as the filter, is altered by the sanitization process, then the permeability of the mask would change.

FFP2 masks, and their US equivalent N95, are required to meet certain standards of filtering. These standards are not directly defined by the permeability of the filter, but on the ability of the filter to stop aerosol when subjected to a certain airflow. The study described herein did not assess aerosol filtering, and therefore does not guarantee that the FFP2 mask will perform as expected regarding aerosol filtering ability. However, aerosol filtering is the consequence of two effects, the permeability of the filter, which we show here remains unchanged, and the wettability of the fibers, which we argue below should not be altered by ethanol. Furthermore we have tested masks from only one manufacturer; and the sanitization protocol employed in this study (1 hour soaking in 99% grade ethanol) has not been optimized to eradicate microbes and viruses.

For comparison, we also measured the permeability of a “homemade” fabric mask meeting the French standard AFNOR (3) and found the mask 4 to 8 times more permeable (offering less resistance to flow, therefore less filtering) than a FFP2 mask.

## Materials and Methods

A permeability measurement was initially performed on a 2 cm^2^ section of a FFP2 mask. The mask was then sanitized in 99% ethyl alcohol and, once the alcohol had evaporated (a couple of hours), the measurement was reproduced on the same section of the mask. This protocol was repeated for six cleaning cycles.

### FFP2 mask

The FFP2 mask (Figure 1) is a mechanical filtering facepiece respirator designed to protect against air particulates such as droplets and aerosols. Its performance standards are defined in the UNE EN 149:2001 “Respiratory protective devices - Filtering half masks to protect against particles - Requirements, testing, marking”. To meet the FFP2 standard, a mask needs to be able to separate at least 94% of airborne aerosols that pass through the filter at 95 L/min air flow. Other requirements in the EN 149–2001 standard, relative to the pressure drop under certain airflow, are directly related to the permeability of the filter.

**Figure 1:**
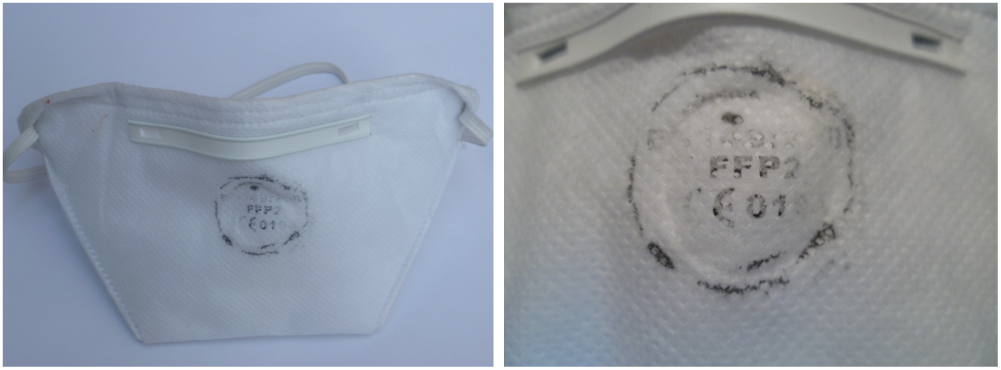
A FFP2 mask. India ink was used to assure that the permeability measurement was performed at the same location.

The European FFP2 masks have very similar filtering capabilities to the American N95 masks. The N95 mask corresponds to the US NIOSH-42C FR84 standard, and must be able to filter at least 95% of aerosols at a flow rate of 85 L/min. Note that we have not performed measurements on N95 masks.

### AFNOR mask

For comparison purposes, we performed an additional measurement on a barrier mask meeting the French standard AFNOR SPEC S76–001 (3). These masks are designed for the general population. Their conception differs greatly, but they are usually made of several layers of fabrics.

### Cleaning procedure

After the initial measurement, the mask was soaked in 99% ethanol (ethyl alcohol 99% grade, purchased from MonDroguiste.com) for 1 hour, and then left at room temperature to allow complete drying and evaporation of the alcohol. No shear was applied to the mask during the sanitization protocol. A permeability measurement was performed and the entire procedure was repeated up to six times.

### Permeability measurements

#### Darcy’s Law

The fluid flow equation in a porous media was first described by French engineer Henry Darcy in 1856 (4), and is usually referred to as Darcy’s law:

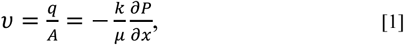

with: *ν* the fluid velocity in the direction x, q the flow rate, A the cross-sectional area, *k* the permeability, *μ* the viscosity of the fluid, and ∂*P/*∂*x* the pressure gradient in the direction x. The permeability has the dimension of a surface area, and is usually expressed in the unit of Darcy: 1 Darcy is equivalent to 9.9×10^−13^ m^2^ or 0.99 μm^2^ (5). Results are also expressed in L / (min mbar cm^2^), which corresponds to a “flow resistivity” and do not depend on the thickness of the measured area.

#### Experimental setup

Permeability measurements were performed using a custom-designed flow permeameter (Figure 2). A 2 cm^2^ section of a FFP2 mask was compressed between a flat rubber seal (5 mm height) and a ceramic plate using a press. The pressure applied by the press was adjusted to limit side leaks. The thickness of the mask was approximately 1 mm. Airflow was regulated by an air pump, and the fluid used was air. Outlet pressure was atmospheric pressure. Pressures in the inlet and the flow rates were recorded using a 16-bit acquisition board and the software CYDAR (6). The experimental set-up was similar to the one described in Lenormand et al. (7).

**Figure 2:**
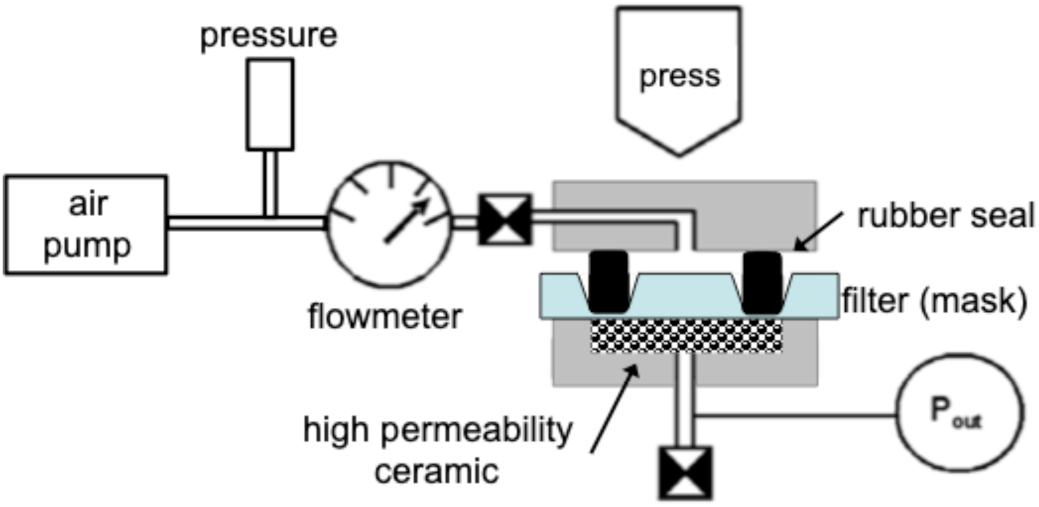
Experimental setup of the permeameter. A 2 cm^2^ section of a FFP2 mask was positioned between a flat rubber seal and a ceramic plate by a press. Pressures and flow rates were measured using the software CYDAR.

#### Permeability measurements and corrections

To measure the permeability of the mask, the pressure gradient was recorded as a function of the flow rate. For each measurement, the flow rate was increased from 0 to 1.2 L/min. This flow rate was applied to a 2 cm^2^ area of the mask and approximated 95 L/min applied to the entire surface of the mask (~180 cm^2^). A measurement typically lasted 10 minutes.

Once data were acquired, the “Permeability” module of the software CYDAR (6) was used to calculate the permeability. The compressibility of air was taken into account in the analysis.

Due to the high permeability of the mask, the pressure drop across the mask was on the same order as the pressure drop in the tubing and end-pieces of the permeameter. Therefore, we initially performed a pressure vs. flow rate measurement of the apparatus, without the mask in place. A second measurement was then performed with the mask in place. The pressure drop due to the mask was calculated as the difference between both curves (Figure 3). The pressure curve without mask (and consequently also the raw data) follows a quadratic equation, due to inertial effects. However the corrected curve is a linear function and therefore follows Darcy’s law (Equation 1). At maximum flow rate, the Reynolds number was estimated to be Re = 0.016. The Reynolds number in the filter was small compared to 1, and the flow is laminar with no inertial effects.

**Figure 3:**
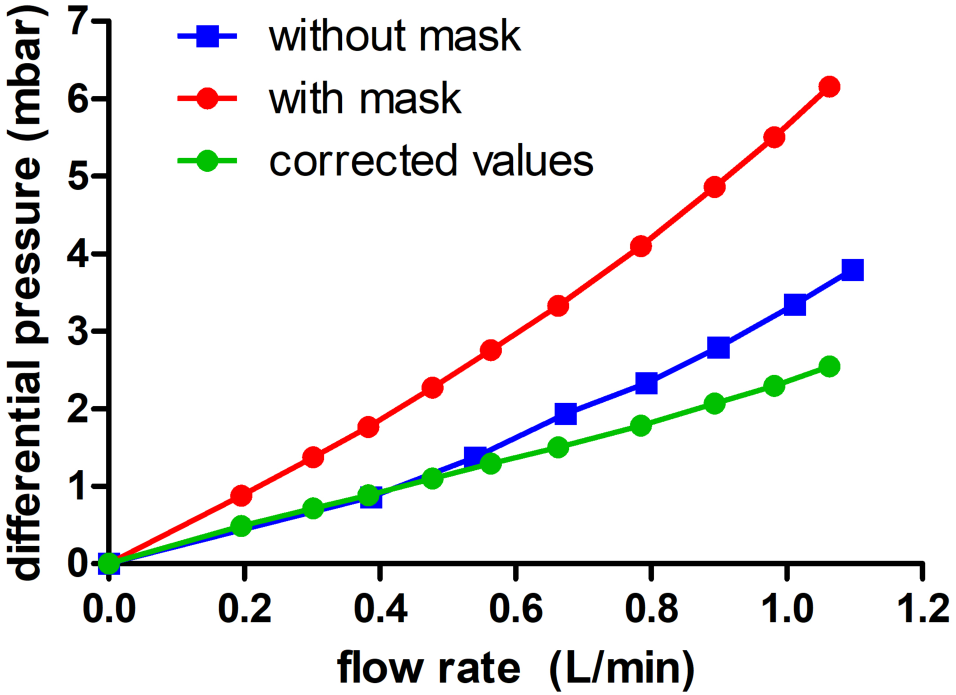
The pressure drop across the mask (green) was inferred from the measurement without the mask (blue) and the measurement with the mask in place (red). The corrected curve (green) is a linear function and follows Darcy’s law (Equation 1).

## Results and Discussion

### Pressure drop measurement

A measurement of the pressure drop as a function of the flow rate is shown in Figure 4. Measurements before cleaning (black circles) and after 1, 2, 3, 4, and 6 cleanings are shown. All measurements appear to be linear functions of the flow rate, as expected from Darcy’s law (Equation 1). Importantly no systematic trend from one measurement to the next was observed.

**Figure 4:**
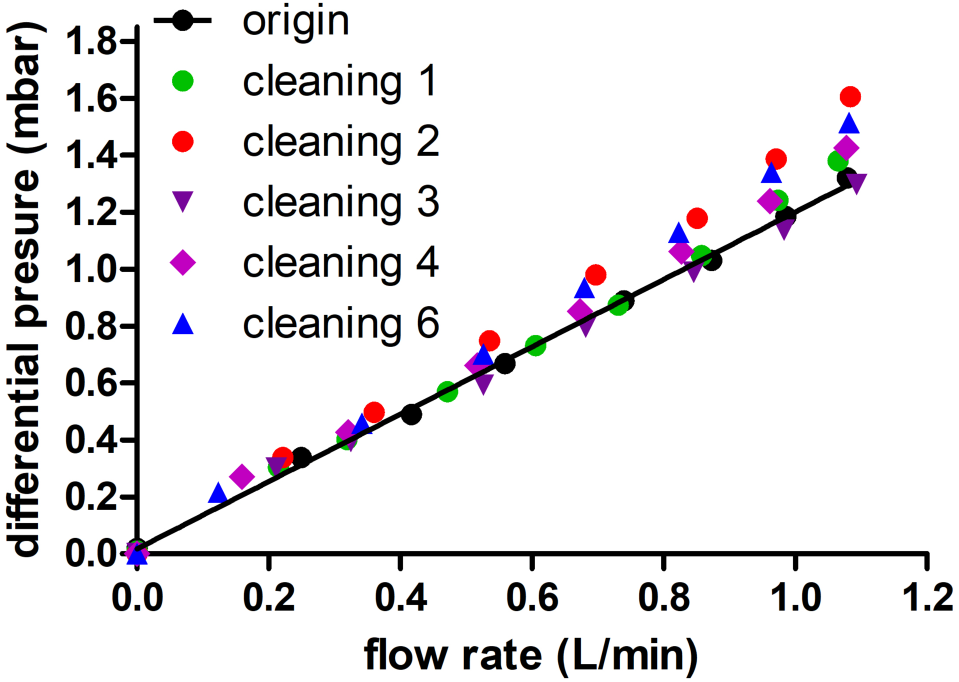
Pressure drop as a function of flow rate before (origin) and up to 6 cleanings on a 2 cm^2^ section of a FFP2 mask. No systematic changes were observed. Error bars in pressure are ± 0.02 mbar; error bars in flow rate are ± 0.01 L/min.

### Permeability

From each measurement curve, the permeability of the calculated from Darcy’s law (Equation 1). For a FFP2 permeabilities were between 6 and 12.5 Darcy, or 0.4 L / (min mbar cm^2^ ). Permeability measurements did significant changes with the number of cleanings (Figure 5) cleaning cycles, the permeability remained very close to the permeability before cleaning; the standard deviations between all measurements were 0.66 Darcy for disk 1 and 0.80 Darcy for disk 2. Differences between measurements could arise from the difficulty of repositioning the rubber seal at the exact location from one measurement to the next. Also, measurements on disk 1 and disk 2 were performed on the same mask, showing the spatial heterogeneities of the filtering media. The variability in permeability resulting from one measurement to the next was less than the variability arising from different area of the mask.

**Figure 5:**
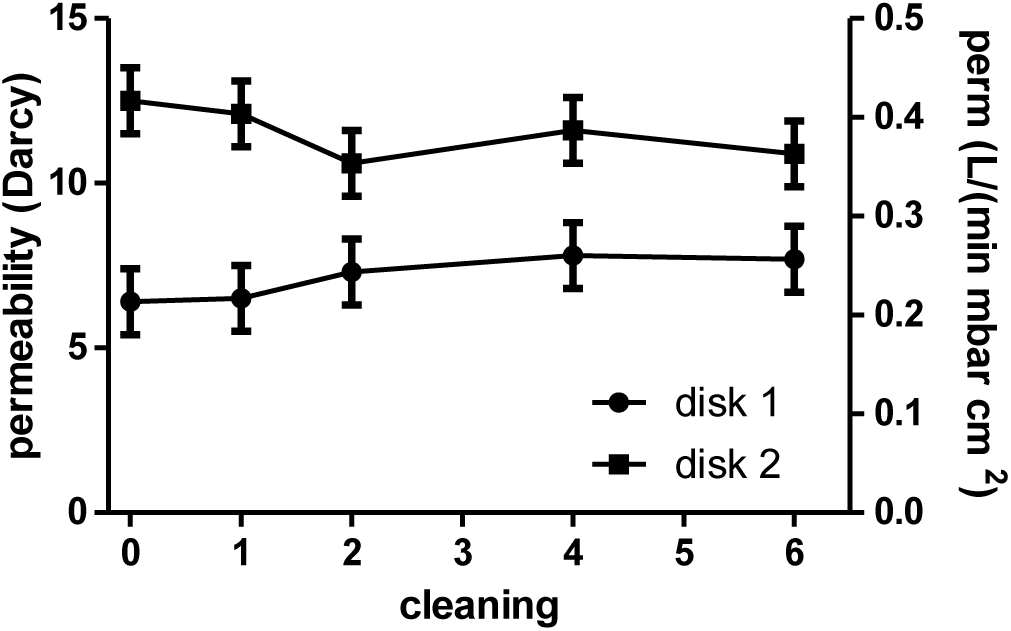
Permeability for two disks on a FFP2 mask derived from each measurement as a function of the number of cleaning. 0 cleaning corresponds to the initial value. No significant change in the permeability was observed.

Measurements performed on the barrier mask showed a permeability of 48 Darcy, or 1.6 L / (min mbar cm^2^). The permeability was 8 times higher than that of disk 1, showing a much lower resistance to airflow, and therefore less filtering capability.

### Limitations of the study

This study suggests that ethanol could be used to sanitize a FFP2 mask without significantly altering its filtering properties.

Measurements were performed on masks from only one manufacturer, therefore it is not established whether or not the results presented here can be extrapolated to masks from different manufacturers.

Furthermore, this study did not assess the aerosol filtration directly, as defined in the EN 149:2001 performance standard. However it is highly probable that the aerosol filtration properties remain unchanged after ethanol cleaning for two reasons. First, the permeability measurements demonstrated that the pore structure of the filter was unchanged by the sanitization protocol. Secondly, ethanol cleaning is not expected to change the wettability of the filter. The filter should remain hydrophilic; water wettability is necessary to retain the aerosol droplets. However, this assumption should be confirmed by a laboratory that is capable of performing the appropriate aerosol tests.

Finally, the sanitization protocol used here, a 1 hour soak in 99% ethanol, was not optimized to eradicate microbes and viruses. We assumed that ethanol would be a likely candidate for such usage, as it is mostly safe and widely available.

The *Centers For Disease Control* proposes several sanitization protocols using ethanol (8). Most of these protocols include a soak for 5 to 10 min in 70% ethanol; or 5 min in 70%-90% isopropyl alcohol.

> *“Ethyl alcohol, at concentrations of 60%–80%, is a potent virucidal agent inactivating all of the lipophilic viruses (e.g., herpes, vaccinia, and influenza virus) and many hydrophilic viruses (e.g., adenovirus, enterovirus, rhinovirus, and rotaviruses but not hepatitis A virus (HAV) or poliovirus). “ (8)*

The protocol in this study used a longer duration and was performed at a higher alcohol concentration, as our main concern was to study alteration to the fibers forming the filter. We do not expect that a treatment at a lesser concentration of ethanol for a shorter time would change the results presented here.

## Concluding remarks

We show here that after six cleaning cycles, the permeability of a FFP2 mask remains very close to the permeability before cleaning. Amid the COVID-19 pandemic and the shortage of protective masks, this study suggests that ethanol could be used to sanitize a FFP2 mask without significantly altering its filtering properties. Additional measurements on masks from different manufacturers need to be performed to validate the results presented here.

## Data Availability

All data presented in this paper can be made available upon request.

## End Matter

### Author Contributions and Notes

R.L. designed and performed research; R.L. and G.L. wrote the software and analyzed data; and G.L. wrote the paper.

The authors declare no conflict of interest.

This article contains supporting information online: http://www.cydarex.fr/files/Cydarex_perm_mask.pdf

## Notes

### Competing Interest Statement

The authors have declared no competing interest.

### Funding Statement

This research was privately funded.

